# Injury of the Corticofugal Tract from the Secondary Motor Area in Middle Cerebral Territory Infarction: A DTI Study

**DOI:** 10.1101/2023.03.02.23286661

**Authors:** Jeong Pyo Seo, Seong Ho Yun

**Author notes:** **Correspondence:** Seong Ho Yun, 119, Dandae-ro, Dongnam-gu, Cheonan-si, Chungcheongnam-do, Republic of Korea.

## Abstract

**Background and Purpose:** Middle cerebral artery (MCA) territory infarction commonly induces a variety of motor function deficits because it involves multiple descending motor pathways, including the corticospinal tract (CST) and corticofugal tract (CFT). Despite the importance of the MCA territory for motor function, there is currently insufficient evidence regarding an injury of the CFT from the secondary motor area in MCA territory infarctions. We investigated injury of the CFT from the secondary motor area and CST in patients with MCA using diffusion tensor tractography (DTT).

**Methods:** Thirty-five patients with MCA territory infarctions and 30 controls were recruited. DTT parameters, including fractional anisotropy (FA) and tract volume (TV), of the CST and CFTs from the dorsal premotor cortex (dPMC) and supplementary motor area (SMA), were analyzed.

**Results:** In the affected hemisphere, the FA values of the CFTs from the secondary motor areas and CST were significantly lower than those in the unaffected hemisphere and control groups. Additionally, the TV of the CFTs from the dPMC and SMA were significantly lower than those from the unaffected hemisphere.

**Conclusion:** We observed concurrent injury to the CFTs from the secondary motor area and CST after MCA territory infarction. Our findings explain the neural mechanisms underlying motor weakness and limb kinetic apraxia in patients with MCA.

## 1. Introduction

The middle cerebral artery (MCA) territory is most frequently affected in ischemic stroke.^1^ It supplies blood to various brain regions, including the frontal lobe, the lateral surface of the parietal and temporal lobes, and integrative associative areas.^2-4^ As the MCA supplies blood to large brain regions, MCA territory infarcts induce a variety of neurological and motor deficits in hand function, posture control, and balance.^5-9^

In the human brain, motor function is associated with various neural tracts, including the corticofugal tract (CFT), corticospinal tract (CST), corticoreticulospinal tract, and corticorubrospinal tract.^10-15^ The primary function of CFTs from the secondary motor area, which is classified as the premotor cortex (PMC) and supplementary motor area (SMA), is motor planning.^16^ Consequently, injury of the CFTs from the secondary motor area commonly induces limb kinetic apraxia, which is the inability to make precise or exact movements with the hands and upper or lower extremities.^17^ Additionally, motor weakness could be associated with CST injury and limb kinetic apraxia, which results from the injury of the CFTs from the secondary motor area.^11^ Therefore, accurate evaluation of CFTs from the secondary motor area and CST states is essential for successful stroke rehabilitation, including prediction of prognosis and establishment of therapeutic strategies.^18-20^

Diffusion tensor tractography (DTT), derived from diffusion tensor imaging (DTI), provides a unique advantage for three-dimensional visualization of CST integrity and estimation.^21,22^ It also allows for three-dimensional reconstruction of the CFTs from the secondary motor area.^10,11,18,23^ Some previous studies demonstrated the injury of CFTs from the secondary motor area in patients with traumatic brain injury, corona radiata infarct, and Parkinson’s disease.^18,23,24^ However, there is currently a lack of evidence regarding limb kinetic apraxia caused by injury of the CFTs from the secondary motor area in patients with MCA territory infarction. Therefore, in this study, we attempted to investigate CFT injury from the secondary motor area and CST in patients with MCA territory infarction using DTT.

## Methods

### Subjects

Thirty-five patients with MCA territory infarct (23 males, 12 females; mean age, 58.2 years; range 34–69) and 30 age- and sex-matched control subjects (20 males, 10 females; mean age, 60.7 years; range 41–80) with no history of neurological or psychiatric disease were recruited in the present study. Patients with MCA territory infarcts were recruited according to the following inclusion criteria: (1) first-ever stroke, (2) age 30–70 years, (3) duration from onset to the time of MRI scanning < 8 weeks, and (4) MCA territory infarct (involving superficial anterior, superficial posterior, and deep territories),^25,26^ as confirmed by a neuroradiologist. Patients who showed severe cognitive problems (Mini-Mental State Examination score < 25) and severe motor weakness (Medical Research Council grade ≤ 2) were excluded. All participants provided written informed consent in accordance with the Declaration of Helsinki. The study protocol was approved by the Institutional Review Board of the Yeungnam University Hospital (YUMC-2021-03-014).

### Diffusion tensor imaging and tractography

DTI data were acquired within 8 weeks of onset using a 1.5 T Philips Gyro Scan Intera (Hoffman-La Roche, Best, The Netherlands) with single-shot echo-planar imaging. For each of the 32 non-collinear diffusion-sensitizing gradients, 65 contiguous slices were acquired parallel to the anterior or posterior commissure line.^27^ The imaging parameters were as follows: acquisition matrix□=□96 × 96, reconstructed to matrix□=□192 × 192, field of view□=□240 × 240 mm, TR□=□10,398 ms, TE□=□72 ms, parallel imaging reduction factor (SENSE factor)□=□2, EPI factor□=□59, *b*□=□1000 s/mm^2^, NEX□=□1, slice gap□=□0, and slice thickness□=□2.5 mm.^27^

The Oxford Centre for Functional Magnetic Resonance Imaging of the Brain Software Library (FSL; www.fmrib.ox.ac.uk/fsl) (FSL version 5.0, FMRIB, Oxford, UK) was used to analyze the diffusion-weighted imaging data. Affine multiscale two-dimensional registration was applied to correct the head motion effect and image distortion due to eddy currents. Fiber tracking using probabilistic tractography in the default tractography method based on a multifiber model was applied with tractography routines implemented in FMRIB (5000 streamline samples, 0.5 mm step lengths, curvature thresholds = 0.2).

CFTs from the dorsal premotor cortex (dPMC), SMA, and CST were determined by selecting the fibers passing through the seed region and target regions of interest (ROI). To reconstruct the CFTs from the dPMC and SMA, the seed ROIs were placed on the crus cerebri on the fractional anisotropy (FA) map, and the target ROIs were placed on the dPMC and SMA, as reported in previous studies.^28,29^ For analysis of the CSTs from the precentral knob, the seed ROI was placed on the CST portion of the lower pons on the color map, and the target ROI was placed on the M1 at the axial slice.^30,31^ The results were visualized at the threshold of two streamlines through each voxel to analyze 5000 samples generated from each seed voxel. The FA and tract volume (TV) of the CFTs and CST were measured.

### Statistical analysis

Statistical analysis was performed using the SPSS software (version 25.0; IBM Corp., Armonk, NY, USA). The Shapiro–Wilk test was used for normality testing among the measurements. Independent t-tests (age) and chi-square tests (sex) were performed to analyze the general characteristics between patients and the control group, setting the level of significance at *p* <0.05. One-way analysis of variance was used to determine differences in DTI parameters (FA and TV) of the CFTs and CST between the patient and control groups. Post-hoc analysis was performed using Bonferroni correction with a level of significance of *p* <0.017.

## Results

DTI analysis in the MCA patient group was performed in 28 of 35 patients. For some patients, CFTs from the secondary motor area (n=4) and CST from M1 (n=3) were not reconstructed because of severe brain injury and Wallerian degeneration. Fourteen of the 28 patients had a lesion in the right hemisphere, while 14 had a lesion located in the left hemisphere.

Table 1 shows the mean values of the DTI parameters of the CFTs from the secondary motor area and the CST in the patient (affected and unaffected hemispheres) and control groups. In the CFT from the dPMC, there were significant differences in the mean FA and TV values between the affected and unaffected hemispheres and the control group (*P*<0.05). The post-hoc analysis revealed that the mean FA values of the affected hemisphere were significantly lower than those of the unaffected hemisphere and control group (*P*<0.05). Additionally, the mean TV value of the affected hemisphere was significantly lower than that of the unaffected hemisphere (*P*<0.05).

**Table 1.**
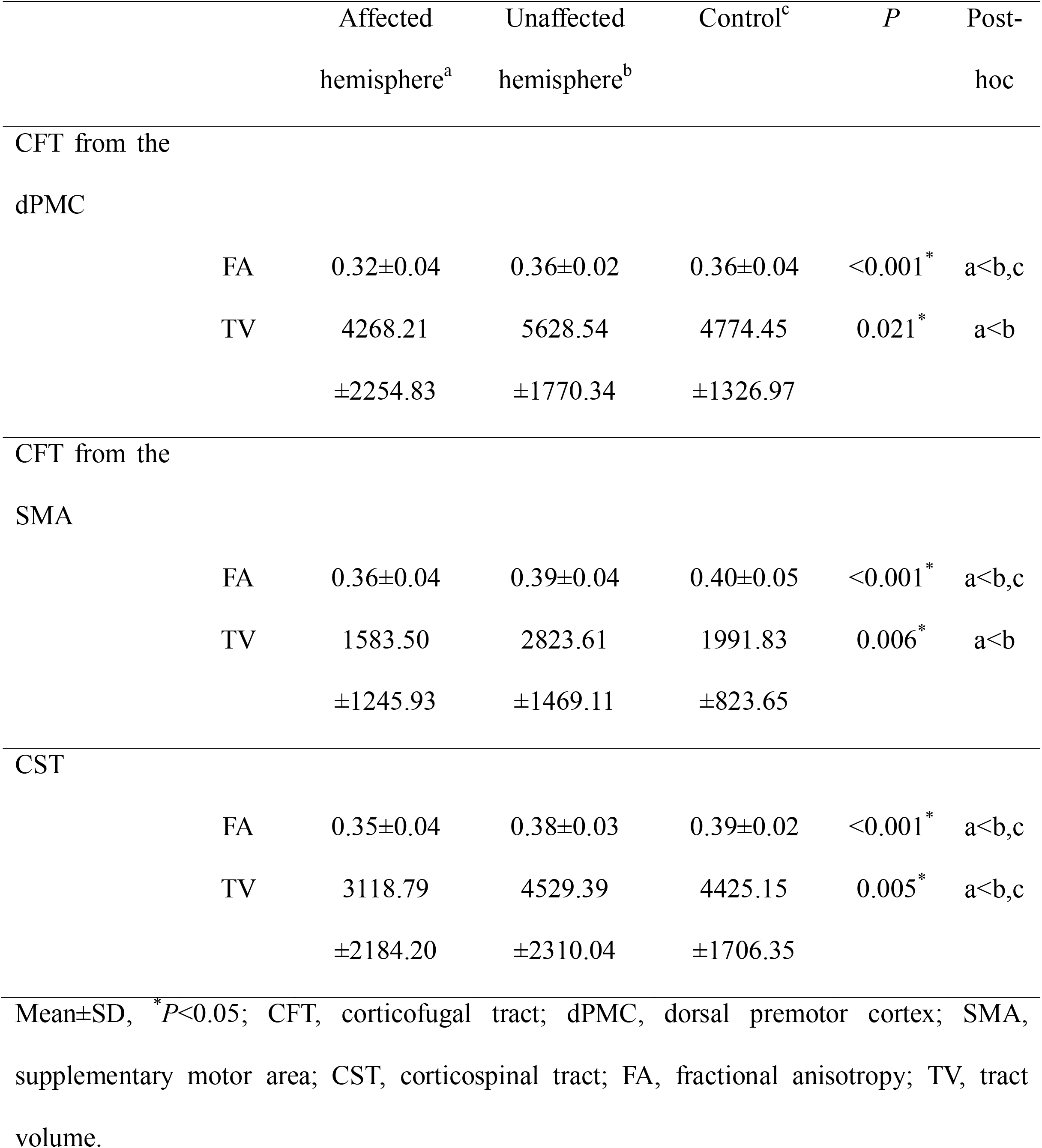
Comparisons of diffusion tensor image parameters of the CFTs from the secondary motor area and CST between patients and the control group

**Figure 1.**
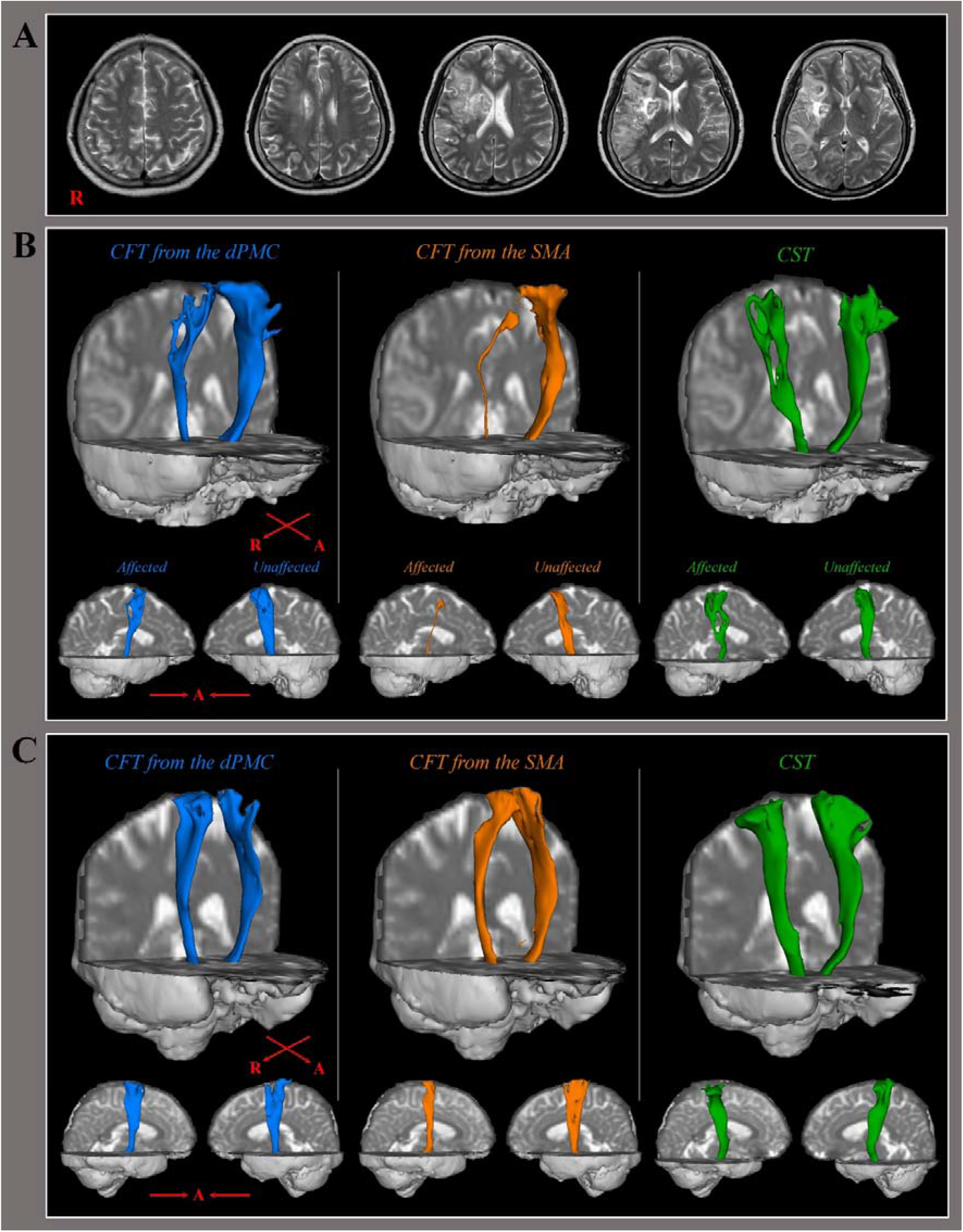
Diffusion tensor tractography (DTT) of the corticofugal tract (CFT) and corticospinal tract (CST) in middle cerebral artery (MCA) infarct patients and age-matched control subjects. A: T2-weighted magnetic resonance images of a patient with right MCA infarct (50s female), B: DTT for the CFT from the dorsal premotor cortex (dPMC) (blue color) and supplementary motor area (SMA) (orange color) and the CST (green color), C: DTT for the CFT from the dPMC and SMA and the CST in a normal 40s female.

Regarding the CFT from the SMA, there were significant differences in the mean FA and TV values between the affected and unaffected hemispheres and the control group (*P*<0.05). The post-hoc analysis revealed that the mean FA value of the affected hemisphere was significantly lower than that of the unaffected hemisphere and the control group (*P*<0.05). Additionally, the mean TV values of the affected hemisphere were significantly lower than those of the unaffected hemisphere (*P*<0.05).

The CST from M1 showed a significant difference in the mean FA and TV values between the affected and unaffected hemispheres and the control group (*P*<0.05). The mean FA and TV values of the affected hemisphere were significantly lower than those of the unaffected hemisphere and control group in the post-hoc analysis (*P*<0.05).

## Discussion

In the present study, we investigated injury of the CFT from the secondary motor area and CST in patients with MCA territory infarction using DTT. In the affected hemisphere, the FA value of the CST and CFT from the dPMC and SMA were significantly lower than those of the unaffected hemisphere and the control group. The TV value of the CST and CFT from the dPMC and SMA were significantly lower in the affected hemisphere than in the unaffected hemisphere. Regarding the DTT parameters, the FA value represents the degree of directionality of water molecule diffusion and integrity of white matter microstructures such as axons, myelin, and microtubules; therefore, it reflects the fiber density, axonal diameter, and white matter myelination.^21,32^ The TV value indicates the total number of neural fibers in the neural tract determined by the total number of voxels.^32^ Consequently, decreases in the FA and TV values of the CST and CFTs from the secondary motor area indicated injury of the neural tracts following MCA territory infarction.

Our results confirmed that MCA territory infarction induces a decrease in fiber integrity and amounts of CST and CFT from the dPMC in the affected hemisphere. The MCA perfused various brain regions, including subcortical regions (e.g., corona radiata, internal capsule, globus pallidus, caudate nucleus, thalamus) and the cerebral cortex (e.g., primary motor cortex and premotor cortex).^33-36^ According to fMRI studies, subcortical infarction in the corona radiata and internal capsule is confirmed following MCA territory infarction.^37-39^ Given that the CST and CFT axons project to the corona radiata and internal capsule,^34,35^ we thought that degeneration of the CST and CFT could be attributed to damage in the subcortical white matter by MCA territory infarction. However, our results showed no significant difference in the TV of the CFT from the dPMC between the affected and control groups. In the acute phase of cerebral infarction, extensive injury of the white matter occurs along with inflammation and edema around the periphery of the hematoma.^40,41^ The CFT from the secondary motor area in some patients with acute MCA was not reconstructed due to severe brain injury. Because we excluded data that could not be reconstructed, it is possible that there was no significant difference between the affected and control groups in the TV of the CFT from the secondary motor area.

It is generally known that the SMA is supplied with blood by the anterior cerebral artery.^42,43^ However, our results indicated deformation of the CFT from the SMA in the affected hemisphere following MCA territory infarction. Considering the CFT fiber projected corona radiata, internal capsule, and cerebral peduncle, which supply blood flow to the branches of the MCA,^16,18,25,44^ we believe that damage to the CFT from the SMA occurred at the subcortical level by the MCA infarction.

In conclusion, we demonstrated injury to the CFT from the dPMC and SMA in ischemic stroke patients following MCA territory infarction. The results of this study provide an understanding of the pathological mechanisms underlying motor weakness and limb kinetic apraxia in patients with MCA. Additionally, detecting injury to the CFT from the dPMC and SMA would benefit the prognosis and planning of intervention strategies for patients with MCA territory infarction. However, this study has several limitations. First, a relatively small number of patients were recruited. Second, the analysis of fiber tracking is operator-dependent and may underestimate the fibers of the neural tract owing to fiber complexity and the crossing fiber effect.^45,46^ Therefore, further prospective studies should be conducted to address these limitations.

## Data Availability

The data that support the findings of this study are available from the corresponding author upon reasonable request.

## Abbreviations

CST: Corticospinal tract
CFT: Corticofugal tract
DSI: Diffusion spectrum magnetic resonance imaging
DTI: Diffusion tensor imaging
DTT: Diffusion tensor tractography
FA: Fractional anisotropy
MCA: Middle cerebral artery
dPMC: Dorsal premotor cortex
ROI: Region of interest
SMA: Supplementary motor area
TV: Tract volume

## Sources of Funding

This research was supported by the National Research Foundation of Korea (NRF) grant funded by the Korea government (MSIP)(2021R1F1A1064353).

## Disclosures

None

## Notes

### Competing Interest Statement

The authors have declared no competing interest.

### Author Declarations

This study was admitted by the insitutional review board of Yeungnam University Hospital (YUMC 2021-03-014)

